# Contrast Response Function Estimation with Nonparametric Bayesian Active Learning

**DOI:** 10.1101/2023.05.11.23289869

**Authors:** Dom CP Marticorena, Quinn Wai Wong, Jake Browning, Ken Wilbur, Samyukta Jayakumar, Pinakin Davey, Aaron R. Seitz, Jacob R. Gardner, Dennis L. Barbour

## Abstract

Multidimensional psychometric functions can typically be estimated nonparametrically for greater accuracy or parametrically for greater efficiency. By recasting the estimation problem from regression to classification, however, powerful machine learning tools can be leveraged to provide an adjustable balance between accuracy and efficiency. Contrast Sensitivity Functions (CSFs) are behaviorally estimated curves that provide insight into both peripheral and central visual function. Because estimation can be impractically long, current clinical workflows must make compromises such as limited sampling across spatial frequency or strong assumptions on CSF shape. This paper describes the development of the Machine Learning Contrast Response Function (MLCRF) estimator, which quantifies the expected probability of success in performing a contrast detection or discrimination task. A machine learning CSF can then be derived from the MLCRF. Using simulated eyes created from canonical CSF curves and actual human contrast response data, the accuracy and efficiency of the MLCSF was evaluated in order to determine its potential utility for research and clinical applications. With stimuli selected randomly, the MLCSF estimator converged slowly toward ground truth. With optimal stimulus selection via Bayesian active learning, convergence was nearly an order of magnitude faster, requiring only tens of stimuli to achieve reasonable estimates. Inclusion of an informative prior provided no consistent advantage to the estimator as configured. MLCSF achieved efficiencies on par with quickCSF, a conventional parametric estimator, but with systematically higher accuracy. Because MLCSF design allows accuracy to be traded off against efficiency, it should be explored further to uncover its full potential.

**Precis:** Machine learning classifiers enable accurate and efficient contrast sensitivity function estimation with item-level prediction for individual eyes.

## Introduction

Visual contrast sensitivity reflects both peripheral and central visual processing ability. As such, it is useful for diagnosing a variety of visual disorders. The simplest, easiest, cheapest and most portable way to quantify this ability is by querying directly—delivering appropriate visual stimuli and recording the resulting behavioral responses. Even with steady advances in quantifying physiological biomarkers of the retina and brain (Calderone et al., 2013; Yarmohammadi et al., 2016), no alternate diagnostic pathway is foreseen that will completely supplant psychophysical testing for evaluating visual system function.

As with all psychophysical tests, however, estimating contrast sensitivity functions (CSFs) requires serial behavioral data acquisition to estimate latent variables, leading to impractically long acquisition times due to high variance in the underlying physiological processes. While full CSFs can therefore have significant clinical value, rapid psychophysical screenings that sacrifice quantitative precision are often more desirable for practical reasons.

Methods have been proposed for accelerating CSF testing while preserving sufficient accuracy for clinical decision making (Gu et al., 2016; Lesmes et al., 2010; Wang et al., 2016). Because of intrinsic noisiness, speeding up the estimates typically requires making assumptions—incompletely justified in most cases—about some model parameters in order to reduce the degrees of freedom of the model to be learned (Treutwein & Strasburger, 1999). This process results in less flexible models that might work well on average but fail to capture details of some individual curves, especially local nuances such as notches (Tahir et al., 2009; Woods et al., 1996).

For example, a common modern assumption about the shape of the CSF exploited in some of the most advanced CSF estimators is that it adheres to a truncated parabola (Zhao et al., 2021). While this shape visually reflects common CSF trends and has been shown to lead to more accurate estimates compared to alternate parametric forms (Watson & Ahumada, 2005), the parameters of the model do not provide mechanistic insight into the underlying physiological construct. Reductionist modeling in this case, therefore, appears to primarily be useful for achieving sufficient test accuracy with fewer data samples, allowing for more rapid tests. Reducing the resulting model to a small number of parameters does provide a useful shorthand for researchers and clinicians to conceptualize and communicate general CSF shape.

The continued expansion of machine learning success over the past two decades can be at least partially attributed to improved ability to constrain highly flexible models by discerning complex patterns in data (Basha & Rajput, 2019). While some of the most prominent successes involve extremely large data sets (Zhou et al., 2017), similar advances can also be applied to much smaller data sets of various types (Kokol et al., 2021). If more flexible CSF models can be trained in reasonable amounts of time, then more appropriate estimates can be achieved for unusual phenotypes not fit well by a traditional, assumption-laden functional form.

Exactly this capability has been achieved with the threshold audiogram, an auditory analog of the CSF (Cox & de Vries, 2021; Schlittenlacher et al., 2018; Song et al., 2015), as well as threshold perimetry tests (Chesley & Barbour, 2020). The current study summarizes the application of similar principles to construct a CSF estimator, along with a preliminary assessment of estimator performance. The result is a highly flexible nonparametric estimator that has the potential to capture a wide variety of individual CSF curves in practical amounts of time. Perhaps most importantly, this single estimator can be tuned toward either efficiency or accuracy, depending on the application or testing time available.

## Modeling Framework

### Background

The contrast sensitivity function is defined as the performance threshold of a behavioral task in response to manipulations of the spatial frequency and visual contrast of a visual stimulus (Ginsburg, 2003). Other stimulus parameters such as mean luminance, size and visual eccentricity are typically held constant or manipulated in stepwise fashion, resulting in a series of CSF curves in the latter case (Kolb et al., 1995). The CSF curve reflects a particular success probability contour on the underlying multidimensional psychometric function, or psychometric field. When considering only a single spatial frequency, signal detection theory predicts that the ability to detect the presence of a visual pattern is a monotonically increasing function of contrast (Green & Swets, 1966; Kingdom & Prins, 2010). The resulting one-dimensional psychometric function *ψ*(*κ*) is modeled as a sigmoid in contrast *κ*.

Because the entire psychometric function is rarely of interest in clinical applications, adaptive staircase methods have long been adopted to estimate only the threshold of this sigmoid (King-Smith, 1984; Levitt, 1971; Treutwein, 1995). Such methods exploit assumptions made about the spread (or equivalently, slope) of the sigmoid in order to set appropriate step sizes that probe either side of the threshold. While fast, these nonparametric methods are inherently unscalable to higher dimensions (i.e., adding additional independent variables that affect the psychometric field) because they offer no efficiency gain with increasing dimensionality. In other words, they have little capacity to incorporate knowledge gained at one combination of independent variables to improve the threshold estimate at another combination. The result is a highly flexible but relatively inefficient CSF estimation method. Such methods have, however, been shown to exhibit superior estimation accuracy relative to parametric methods (Watson & Ahumada, 2005).

Incorporating the psychometric spread into a psychometric curve estimator as a free parameter is possible (Leek, 2001), though this typically requires about an order of magnitude more data than when estimating threshold alone (Kingdom & Prins, 2010; King-Smith & Rose, 1997; Kontsevich & Tyler, 1999). These methods are scalable to multiple dimensions (DiMattina, 2015; Lesmes et al., 2006), but the gains to efficiency by sharing inference across all the input variables is generally negated by the additional data requirements to estimate the full function. Therefore, estimating a full psychometric field of contrast responses by combining a parametric estimate of psychometric spread with a low-order parameterization across spatial frequency would result in a relatively inflexible estimator suffering from poor accuracy and efficiency relative to alternatives. Likely for these reasons, no estimator of this type is known to have been developed for CSF estimation.

Because the CSF is a threshold curve, parameterizing it directly and making appropriate assumptions about psychometric spread represents a reasonable compromise to gain sufficient accuracy and practical efficiency. Indeed, multiple parametric forms of the CSF have been evaluated for accuracy and efficiency (Rohaly & Owsley, 1993; Watson & Ahumada, 2005). A balance between model complexity and ability to map individual CSFs, along with some interpretability of model parameters, provides the truncated parabola with an advantage over other model forms. These advantages were exploited to develop the time-efficient quickCSF method (Lesmes et al., 2010), as well as modern extensions with even more desirable characteristics (Zhao et al., 2021).

A practical and effective modern computational CSF estimation therefore exists that appears to balance efficiency and flexibility, but there may still be room for improvement. For example, while demonstrably better than other models, the truncated parabola is unable to adequately capture the natural detail of some individual CSFs (Chung & Legge, 2016; Rohaly & Owsley, 1993). Further, despite the interpretability of the truncated parabola parameters regarding the shape of the CSF curve itself, relationships to underlying physiological variables have yet to be established, compromising any potential mechanistic interpretation of the resulting CSF models.

Nonparametric estimators historically exhibit great flexibility but poor efficiency, as described above. The machine learning field, however, has expended enormous efforts over the past 2 decades developing highly flexible estimators for a wide variety of applications. Exploiting such advances may pay dividends for CSF estimation. Indeed, by recasting the psychometric field estimation problem from one of multiple regression to one of probabilistic classification, new machine learning tools can be brought to bear on this longstanding problem. These methods exhibit little apparent advantage for one-dimensional psychometric function estimation, but considerable advantages for multidimensional estimation (Song et al., 2017), particularly when accompanied by optimal task item selection in the form of Bayesian active learning (Song et al., 2018).

This study involves developing a probabilistic machine learning classifier for estimating a fully predictive contrast response function (CRF), then extracting a CSF estimate from the resulting model. Canonical and real CSF curves are used to create generative models, which then provide simulated behavioral responses representing ground truth values. The ability of the new estimator to fit these known functions is evaluated for different stimulus presentation sequences and bias conditions. The performance of the algorithm is evaluated in terms of accuracy and efficiency and compared against quickCSF performance on the same simulated eyes.

### Gaussian Process Classification

A full development of the following can be found in (Song et al., 2017). Briefly, define *f*(**x**) to be a latent function defined on a continuous multidimensional space **x** ∈ χ. For the current application, **x** represents an ordered pair (ω, *κ*) indexing a visual grating at a particular spatial frequency and contrast. The latent function itself represents the probability of correctly detecting this grating. A Gaussian process (GP) represents a convenient means to encode prior knowledge about the latent function:

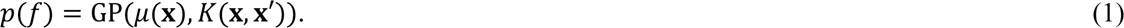

In other words, a GP provides probabilistic constraints on what form the latent function can take. The knowledge represented by the GP can be updated following new data collection according to Bayesian principles. We evaluate the GP over a finite sample of independent values **X** = {**x**_**1**_, **x**_**2**_, . . ., **x***_n_*}. In binary classification tasks the dependent variable can take on one of two values indicating failure or success: *y_i_* ∈ {0,1}. The probability of success *p*(*y* = 1|*f*) is modeled by a sigmoidal link function *ψ*(**x**). This function is distributed according to a Bernoulli likelihood:

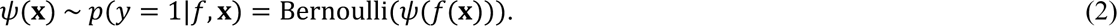

The general sigmoidal link function is given by

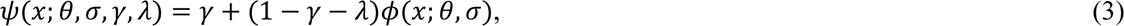

where *ϕ*(·) is the normal cumulative distribution function (CDF) parameterized by threshold (i.e., mean) *θ* and spread (i.e., standard deviation) σ. Remaining parameters include the guess rate *γ*, and the lapse rate *λ* (Wichmann & Hill, 2001). The threshold as a function of spatial frequency *θ*(ω) becomes the CSF. The guess rate for a two-alternative forced-choice task, for example, would be set to 0.5 and for a detection task might be set to 0. The current study simulated only detection tasks and fixed *λ* = *γ* = 0.04 to allow for a small proportion of lucky guesses and lapses of attention. Incorporating any reasonable response error rate value into a detection task estimator tends to improve estimation accuracy relative to an error rate of 0 when errors actually do occur. However, the impact of the precise value selected was not investigated in the current study. Individualized threshold and spread functions *θ*(ω) and σ(ω) were learned by the algorithm for each person.

The resulting machine learning contrast response function (MLCRF) estimator implements a multidimensional nonparametric probabilistic classifier subdividing the input domain of the psychometric field into subdomains of “task success” and “task failure.” Reframing the modeling problem in terms of classification allows algorithmic advances in machine learning classification to be exploited. The CSF reflects a single fixed-probability contour on the full MLCRF, giving rise to the MLCSF as our functional estimate comparable to other CSF estimation algorithm outputs.

### Bayesian Active Learning

A full development of the following can be found in (Song et al., 2018). Briefly, Bayes’ rule is applied to compute an updated GP posterior upon observation of data points {**y**, **X**} from one eye. The posterior is a GP and probabilistically constrains the latent function as described above. Because of the nonlinear link function and use of the Bernoulli likelihood, the posteriors cannot be solved for in closed form. They are therefore estimated via approximate inference techniques, in this case, variational inference (Hensman et al., 2015; Titsias, 2009).

Variational inference is a technique that finds the best approximation of the true posterior distribution from a family of simpler distributions by minimizing the Kullback-Liebler divergence between the approximate and true posteriors (Gardner et al., 2018). We can efficiently estimate the posterior distribution of the GP model, which when trained with all existing data for that eye can compute model updates for any new sample **x**^∗^ ∈ **X**^∗^ defined over spatial frequency and visual contrast. Therefore, the new sample **x**^∗^ that, upon observation, maximizes some utility function *U*(**x**^∗^) is optimal. We define an acquisition function for obtaining this sample as

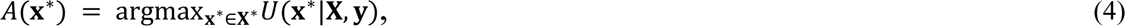

where *U*(·) reflects model quality. We implement uncertainty sampling by defining the utility function as the differential entropy, which quantifies the uncertainty associated with the predictive distribution. In this specific acquisition function, the differential entropy serves as a proxy for information gain, meaning that it aims to select the next sample point **x**^∗^ that would maximize the information about the underlying latent function (Houlsby et al., 2011). This is consistent with the overall goal of Bayesian active learning, which seeks to build an accurate model with as few samples as possible.

## Methods

### Simulations

Ground truth models for four canonical CSF phenotypes were constructed from textbook threshold curves, as depicted in **Figure 1** (Kalloniatis & Luu, 1995). These curves are maximally smooth in shape and occupy extremes in the domain of likely CSFs. Similarly, ground truth CSF models for seven neurotypical individuals and twelve individuals with a diagnosis of schizophrenia were constructed from threshold values extracted at discrete spatial frequencies during a CSF training regimen (**Figure 2**).

**Figure 1:**
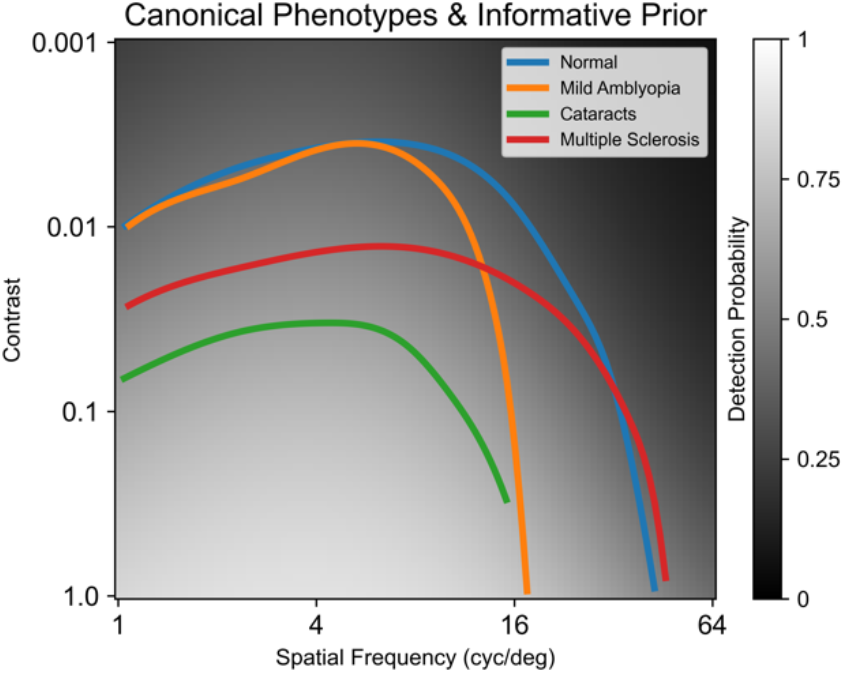
Canonical CSF curves representing typical contrast detection thresholds for four different disease phenotypes. These phenotypes illustrate the general shapes of CSFs as well as variations in loss of visual function across spatial frequency for different clinical conditions. Prior belief used for all informative prior conditions is shown in grayscale.

**Figure 2:**
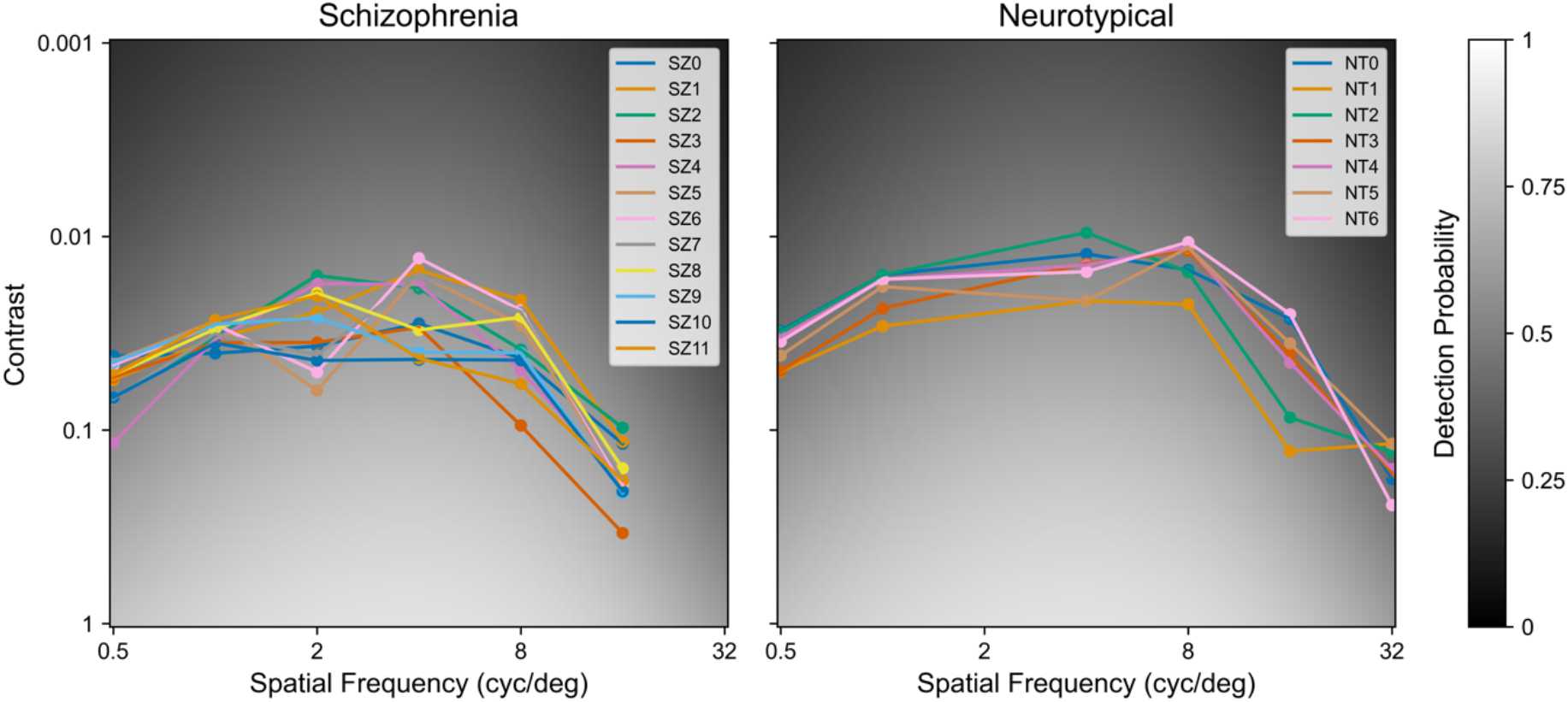
CSF curves estimated from an intermediate stage of a contrast training study for participants with schizophrenia (left) and participants with no known neurological disorder (right). These CSFs have more local variations than the overly smooth canonical curves. Prior belief used for all informative prior conditions is shown in grayscale.

The convention for all plots similar to Figures 1 and 2 is logarithmic spatial frequency versus logarithmic visual contrast. Spatial frequency tick marks indicate octaves while contrast tick marks indicate decades. Even CSF curves are plotted on axes of contrast as opposed to reciprocal contrast to promote consistency when responses to stimuli at various contrast values are also plotted on the same axes. Quantification of curve fits are made in units of log contrast.

In all cases, threshold curves as a function of spatial frequency were simulated by splines. The resulting threshold curves were used to create generative ground truth CRF models representing a visual pattern detection task as a function of spatial frequency and visual contrast. At each spatial frequency a one-dimensional logistic psychometric curve was constructed using the four-parameter model given in equation 3 to define the mean, corresponding to the 0.5 probability detection threshold. The sigmoid for the simulations was given by the standard logistic function:

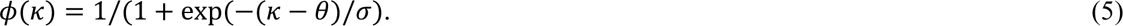

A different sigmoid than that modeled in MLCRF was deliberately selected to evaluate estimator robustness. A plausible spread of 0.08 was simulated across all spatial frequencies (Zhao et al., 2021). The impact of spread on the behavior of this type of estimator across 2 orders of magnitude has been explored previously, and its precise value is unlikely to influence the initial conclusions drawn about estimator performance (Song et al., 2017, 2018). At any indicated contrast level, binary observations (i.e., detected or not detected) were generated by sampling from a Bernoulli distribution with success probability given by *ψ*. A total of 23 generative models formed the collection of simulated eyes evaluated in this study.

### Psychometric Field for Contrast Response

The psychometric field defining the CRF over the domains of spatial frequency and contrast is given by *ψ*(ω, *κ*), while the underlying latent function is defined by *f*(ω, *κ*). With the setup described above, the entire estimation procedure for the CRF is reduced to learning a GP over the latent function. A GP is completely determined by its mean and covariance functions (Rasmussen & Williams, 2006). By selecting closed forms for these functions, estimation further reduces to updating the corresponding parameters, which are then combined with observed data to generate a posterior belief about the CRF. Because the parameters in question reflect the GP rather than the latent function, they represent hyperparameters of the overall model, yielding a formally semiparametric estimator for contrast response.

Many different functional forms can be selected for the mean and covariance functions to incorporate a set of assumptions about the form of the latent function. These form looser constraints than familiar parametric model forms, retaining flexibility to fit a wide variety of functional shapes while potentially fitting those shapes with fewer data than other estimator classes.

The GP mean function is assumed to be constant:

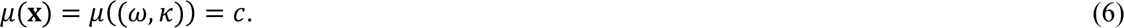

This may seem to be a counterintuitive choice because the shape of the CSF is known not to be flat. It is important to remember, however, that the mean function parameterizes the GP and not the latent function. In this form it is capturing the overall contrast sensitivity across all spatial frequencies. As such, this hyperparameter alone could be useful at distinguishing, for example, cataracts from normal vision.

Covariation along the contrast dimension is assumed to be linear. When combined with the normal CDF link function, the result is a sigmoid that is shifted and scaled to reflect the psychometric properties of contrast detection. Covariation in the spatial frequency dimension is assumed to be continuous and smooth. Parametric estimators cannot easily represent such broad constraints without incorporating many parameters, typically increasing data requirements to fit all of them. The form of the GP covariance function, on the other hand, can be compactly formulated to represent both the linear and smoothness assumptions:

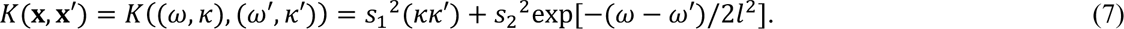

The hyperparameters *s*_)_ and *s*_*_ represent scaling factors and *l* represents a length constant along the spatial frequency dimension. Other kernel designs are possible, but this design has been particularly successful at estimating behavioral functions similar to the CSF (Song et al., 2017).

### Implementation

All simulation, machine learning, and evaluation software was written in Python using major libraries PyTorch (*PyTorch 1.13*, n.d.) and GPyTorch (*GPyTorch 1.8.1*, n.d.). Project information, including code and data necessary to replicate these experiments, can be found at https://osf.io/cpkn5/.

Fifteen logarithmically spaced spatial frequencies per octave were used for the estimation such that ω ∈ [1 64] cycles per degree (i.e., 6 octaves) for experiment 1 and ω ∈ [0.5 32] cycles per degree for experiment 2. Contrasts used for the estimation included *κ* ∈ [0.001 1] (i.e., 3 orders of magnitude) with 30 logarithmically spaced values per decade. Therefore, input values used for modeling, prediction and quantification were discretized over a 91×91 grid with just over 8000 values. This resolution is arbitrary and could be modified as needed.

Samples were generated either via randomly drawing uniformly from all possible combinations of frequency and contrast on the evaluation grid, or from actively learning the most informative next sample for estimation. In both cases, two types of priors were selected to combine with newly observed data to form the posterior. One was uninformative, including no information about CSFs, and the other was informative, including canonical CSF shape information.

For the uninformative prior condition, the mean function of the GP was initialized with *c* = 0. Zero on the latent function maps to a probability of success of 0.5, implying that without any data, the estimator assumes maximum uncertainty about the shape of the CRF. The covariance function was initialized such that *s*_1_ *= s*_2_ *= l =* 1. The intention of this prior was to allow the sampled data to speak for themselves in order to deliver a final estimate with few assumptions. In every condition, a set of phantom shaping data points were added to assist estimator convergence. These values indexed detection failures at locations well beyond any reported human CSF curve. At each octave of spatial frequency, a phantom failure at a contrast of 0.0005 was added. Another phantom failure was added at (128, 1).

For the informative prior condition, 1000 uniformly random samples across spatial frequency and contrast were divided equally among the four canonical phenotypes observed in the range of [0.5 64] cycles per degree and labeled by the corresponding generative models. A single GP was fit over [1 64] cycles per degree to this entire set of observations. The scaling factor of the linear kernel was then multiplied by 0.4 in order to expand the transition between stimulus response regions. This manipulation “flattened the prior” to weaken the bias it injected into the model while still staying informative. The posterior mean of this GP was then used to initialize the mean function of all later GPs. This exact prior was used for experiment 1. For experiment 2, the GP was fit over [0.5 32] cycles per degree. The relationship between this prior and ground truth CSFs can be seen in Figures 1 and 2, revealing a wide transition between behavioral response regions overlapping the threshold boundaries.

The spatial frequency and contrast of the first eight samples for all experimental conditions were deterministically selected according to a Halton set (Halton, 1964). Halton sets are space filling but not random or grid-like, so every condition experienced identical primer sequences that sampled the stimulus space broadly. Subsequent stimuli were selected either randomly or actively. At the expense of some overall efficiency, this procedure promotes stable active learning. The primer sequence can alternatively be selected by other criteria, such as the set of stimuli determined from population screening studies to be most useful for distinguishing important phenotypes.

Two separate experiments were conducted to test the new estimator under different conditions. For experiment 1, four ground truth generative models were created from the four canonical phenotypes depicted in Figure 1 via high-density sampling and cubic spline fitting. Simulated psychometric spreads were fixed at 0.08. These phenotypes were selected to demonstrate estimator performance under extremes of phenotypic variation. Four combinations of sampling methods (random, active) and prior selection (uninformative, informative) were used to acquire data from the generative models. After each new data point, the CRF was updated as a posterior defined over the entire input domain. The *ϕ* = 0.5 contour of the predictive posterior mean of the CRF became the CSF estimate because this value forms the equiprobable boundary between the two response classes. For the symmetric lapse and guess rates used here, this is also equivalent to *ψ* = 0.5. At octaves of spatial frequency relative to 0.5 cycle/degree, the root mean square error (RMSE) in units of log_10_ contrast between the ground truth CSF and the estimated CSF was quantified. Spatial frequencies for which the CSF would have taken values greater than 1 are excluded from this calculation. The estimated CSF was discretized to the nearest contrast grid value. Each phenotype was evaluated separately for 10 repetitions and the average behavior summarized.

Because the canonical examples are overly smooth, experiment 2 made use of generative ground truth models taken from a cohort of neurotypicals and a cohort of individuals diagnosed with schizophrenia performing a contrast detection task as part of a previous study (Yaghoubi et al., 2022). The study was conducted across three sites: Weill Cornell Medicine (WCM), Nathan S. Kline Institute for Psychiatric Research (NKI), and the University of California, Riverside (UCR). Seven neurotypical (NT) participants and 12 patients with schizophrenia (SZ) were recruited for the study. The total number of participants at each site was as follows: WCM: 6 (4 males; age: mean = 33.5 yrs, SD = 8.48); NKI: 6 (3 males; age: mean = 45.6 yrs, SD = 9.54); and UCR: 7 (2 males; age: mean = 19.93 yrs, SD = 2.15). All subjects reported normal or corrected-to-normal vision.

The gamified training paradigm used in this experiment derived from (Deveau et al., 2014). The task was administered using an Apple iPad Pro 12.9 inch screen (second generation) at a luminance of 600 cd/m^2^, resolution of 2732 × 2048 pixels and pixel density of 264 pixels per inch. The viewing distance of all participants from the screen was 20 inches. The iPads used at all sites were calibrated similarly to reduce the variance between each site where the study was conducted. The stimulus set consisted of Gabor patches (targets) at 6 spatial frequencies. An initial test was performed for each group of participants to approximate the maximum spatial frequency that could be perceived. The spatial frequencies used for NT were 0.5, 1, 4, 8, 16 and 32 cycles per degree and for SZ were 0.5, 1, 2, 4, 8 and 16 cycles per degree, respectively. The stimuli were also presented in 8 orientations (0°, 22.5°, 45°, 67.5°, 90°, 112.5°, 135°, 157.5°). Gaussian windows of Gabors varied with σ between 0.25° and 1° and with phases (0°, 45°, 90°, 135°).

Each group of participants underwent a slightly different training procedure, i.e., SZ performed training for up to 40 sessions (1 session per day), with each session lasting approximately 30 minutes. NT performed 40 training sessions in 20 days, i.e., two sessions per day. Each session consisted of different blocks where Gabor patches at all 6 spatial frequencies were presented.

Each block lasted for 120 s where an array of targets with randomly selected orientation and increasing spatial frequency appeared all at once scattered across the screen (**Figure 3**). The contrast of the target was adaptively determined using a 3 down 1 up staircase. Contrast was decreased whenever 80% of the targets were selected and increased when fewer than 40% of the targets were selected with a 2.5 s per target time limit. Staircases were independently run on each spatial frequency across blocks of training. Spatially varying auditory feedback was given to the participants, i.e., low frequency tones corresponded with targets on the bottom of the screen whereas high frequency tones corresponded to stimuli at the top of the screen. Thus, the horizontal and vertical locations on the screen each corresponded to a unique tone. The sounds provided an important cue to the location of the visual stimuli and were included to boost learning as has been found in studies of multisensory facilitation (Shams & Seitz, 2008).

**Figure 3:**
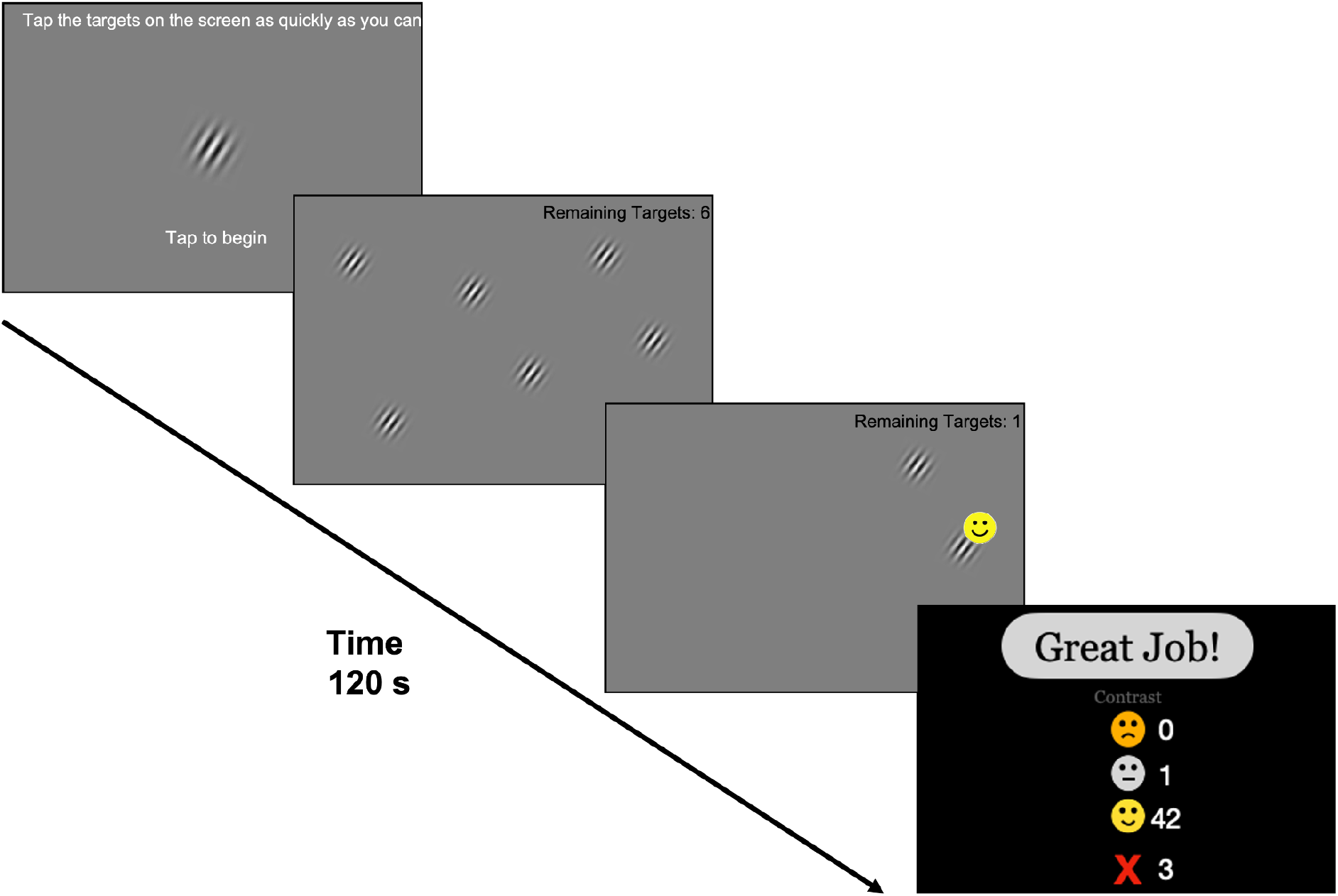
Task design used in experiment 2. Participants were initially presented with a sample target image and were also shown the remaining number of targets that needed to be identified on the screen. At the end of each block, they were provided a summary of their performance. Training blocks varied in terms of the orientation and spatial frequency of the targets.

Like most CSF tests, the CSF values from this study were computed at a small number of discrete spatial frequencies. Spline interpolation was again used to create smooth ground truth CSFs, this time of individual participants’ CSF curves. A value of 0.08 was again used as the fixed psychometric spread to produce simulated contrast response values from the generative models. Newly generated raw data in this fashion were used to train a multidimensional GP probabilistic classifier, as in experiment 1. Estimated CSF values were again compared to ground truth CSF values at octave spatial frequencies with RMSE. Instead of multiple repeats of the same phenotype, however, in this case performance was averaged across all 19 individuals in the data set.

Standard machine learning tuning procedures to achieve consistent model convergence and high model accuracy were conducted only for experiment 1. All estimator configurations for experiment 2 were fixed at these values and not adjusted in an attempt to improve outcomes.

### Evaluation

In all cases the predictive posterior mean over spatial frequency and contrast is taken as the output of the MLCRF estimator. Because of the nature of Gaussian processes, this is equivalent to a *maximum a posteriori* (MAP) estimate. It should be kept in mind, however, that this method is fully Bayesian, meaning that every point estimate in the displayed predictive posteriors actually represents a single point in a complete distribution.

MLCSF estimates were extracted from MLCRFs as the 0.5 probability counter as a function of spatial frequency. This definition has been shown empirically in both simulations and in human experiments to represent an unbiased estimator of the underlying threshold (Song et al., 2015, 2017). Deviation from ground truth was quantified by RMSE as described above.

### QuickCSF Comparison

To discern relative performance of MLCSF against an established CSF estimation procedure, an independent implementation of the quickCSF method (Lesmes et al., 2010) was evaluated under similar experimental conditions. This code base (Canare et al., 2019) was adopted with minimal modification, limited to altering the task design from discrimination to pure detection (i.e., no response errors simulated or estimated) and replacing the source of responses from the default method to the same generative modeling procedures used for MLCSF. Performance was quantized using the same procedures as for MLCSF.

## Results

### Experiment 1

The first experiment involved estimating several canonical CSF phenotypes. **Figure 4** depicts for each canonical phenotype the results of one of these estimation runs with 100 randomly sampled combinations of stimulus spatial frequency and contrast, beginning with an uninformative prior belief. The 0.5 probability contours of the MLCRF predictive posterior mean functions visually correspond to the ground truth CSF functions to a reasonable degree. The capability of this estimator to infer smooth functions indicative of standard disease variants is apparent.

**Figure 4:**
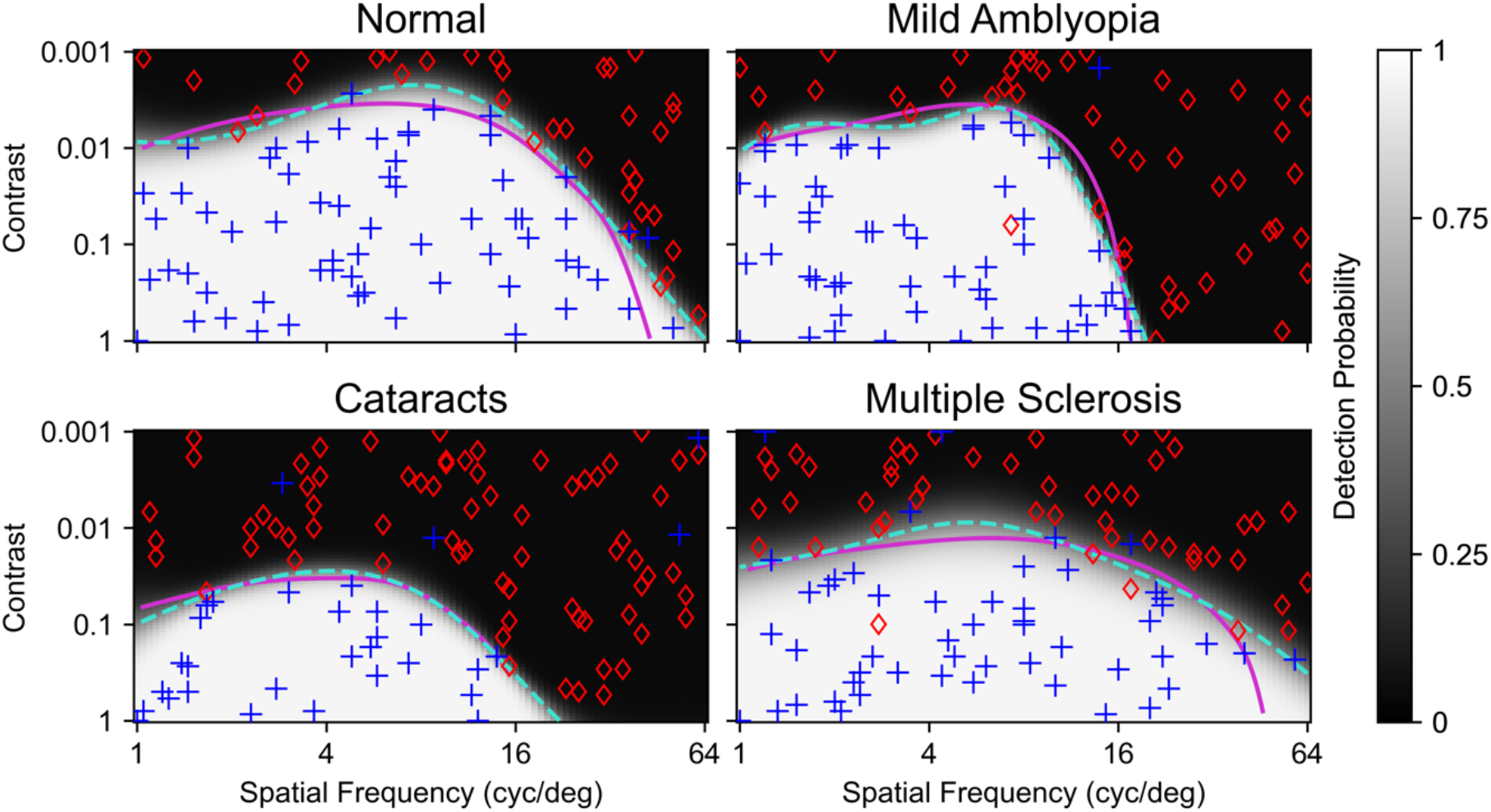
Four canonical CSF phenotypes were converted to generative models in order to create simulated binary detection data. Ground truth CSF curves are shown in magenta. Successful detections of a visual pattern in the stimulus are indicated by blue plusses, while failures are indicated with red diamonds. One hundred samples are randomly distributed in spatial frequency and contrast. Grayscales indicate the learned predictive posterior means from the MLCRF estimator. Dashed turquoise lines represent 0.5 detection probability thresholds determined from the predictive posteriors, indicating that the CSF functions were learned reasonably well. Deviations from ground truth clearly arise from a low density of sampling in the vicinity, which is a limitation of random sampling and fixed-location sampling (i.e., grid search or method of constant stimuli, not shown). Root-mean-square error (RMSE) values in log contrast units for these MLCSF curves are as follows: Normal, 0.205; Mild Amblyopia, 0.139; Cataracts, 0.0678; Multiple Sclerosis, 0.148.

The limitations of random sampling are also on view, as there are multiple regions where, by chance, no samples were taken near threshold. Samples near threshold are intrinsically more informative about the location of that threshold than samples farther away. One of the substantive contributions of modern machine learning has been the development of numerical algorithms for implementing optimization over a wide variety of functional definitions. Even though MLCRF estimates a nonparametric function, for example, the most informative data to collect for refining that estimate can readily be computed within this framework, whereas that is not feasible with classical nonparametric methods.

Active learning is the machine learning principle that seeks to select new samples (i.e., stimulus parameter combinations) such that their observation provides the most information about the latent model. This procedure is analogous to optimal adaptive Bayesian estimation for parametric models (Kontsevich & Tyler, 1999; Watson & Pelli, 1983). An example with 92 samples of active learning (following 8 Halton primer samples) and uninformative prior for the canonical multiple sclerosis phenotype is shown in **Figure 5**.

**Figure 5:**
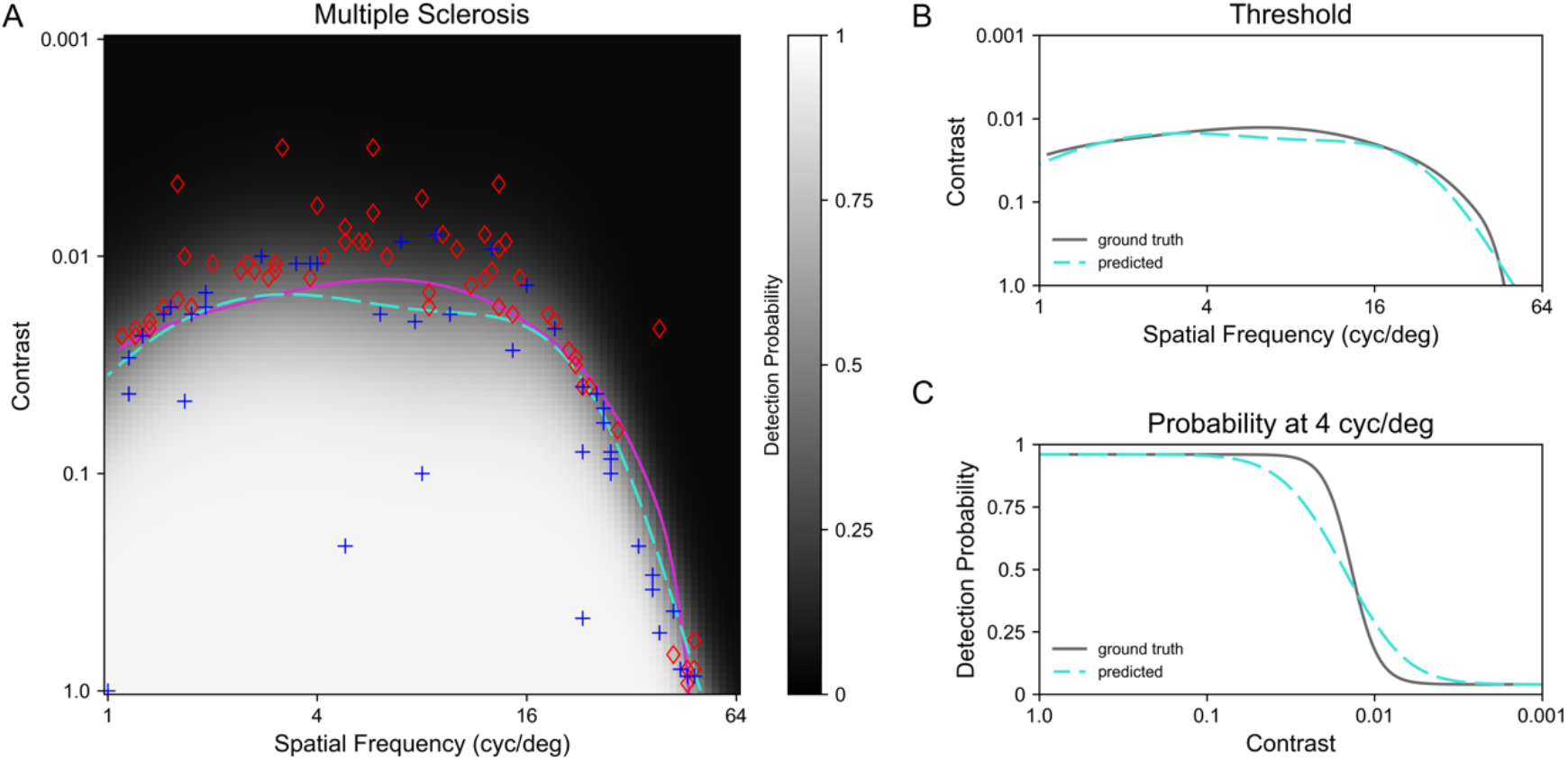
The MLCRF estimated with 100 samples of active learning for one canonical phenotype. **A**. Simulated behavioral responses (blue plusses=success, red diamonds=failure), ground truth multiple sclerosis CSF (solid magenta curve), learned MLCSF (dashed cyan curve) and predictive posterior mean (grayscale) plotted according to the conventions of Figure 4. **B**. MLCSF curve (dashed cyan) replotted along with the ground truth CSF curve (solid). Ordinate units indicate stimulus contrast. **C**. MLCRF psychometric curve at 4 cycles per degree (dashed cyan) plotted along with the ground truth psychometric curve (solid). RMSE value in log contrast units for this MLCSF curve is 0.089.

Active learning involves optimally selecting the next stimulus to deliver based on how informative that new data point would be for updating the model. After training model *M_i_* on the initial *i* sample points during active learning, the most informative next sample point is determined, that stimulus is delivered, the simulated participant response is observed, and a new model *M_i_*_+1_ is learned with *i*+1 points and model *M_i_* as the prior. This procedure continues until data from 100 samples are accumulated.

As can be seen in **Figure 5A**, most of the samples in the active learning condition are distributed near the eventual threshold curve, which would be expected for an acquisition function seeking to maximize informativeness. The ability to rapidly converge on the most informative samples is a key contributing factor of real-time optimization enabling more efficient testing procedures. Most of the samples away from threshold aid in recovering from false positive responses that occurred early in the simulated experiment. The predictive posterior MLCRF mean function can be seen in grayscale, indicating that at every combination of spatial frequency and contrast within the domain of support, the MLCRF estimator returns an estimated probability of stimulus detection. This ability to deliver item-level predictions makes the MLCRF a fully predictive model—in other words, a complete psychometric field.

Direct comparisons between estimates and ground truths for this example are visualized in the side panels. The MLCSF estimate and ground truth threshold as a function of spatial frequency align visually in **Figure 5B**. Accurate estimation of the CSF is the goal of this research because of its established clinical utility. Just as in the case of models employing a truncated parabola, the MLCSF is continuous. Its lack of rigid shape constraints, however, demonstrates its flexibility, indicated here by subtle curvature details.

The MLCSF represents a contour within the MLCRF, made in the current experiments by slicing the predictive posterior mean function parallel to the spatial frequency / contrast plane at a value of 0.5. One can also slice the MLCRF perpendicular to this plane at a single spatial frequency in order to reveal its psychometric shape. The resulting sigmoid for this example at 4 cycles per degree is depicted in **Figure 5C**, showing the learned correspondence to the generative ground truth model. Estimating a single psychometric curve accurately with conventional methods is generally considered to require hundreds of samples, yet only 100 samples total have been accumulated for the entire MLCRF model here. This rapid convergence to a fully predictive model reveals the efficiency of the estimator.

Recall that the MLCRF is a probabilistic classifier, so its predictive posterior reflects a subdivision of feature space into separate regions likely to give rise to one observation or the other (i.e., success or failure). Greater uncertainty of the class boundary leads to wider transitions between regions, which translates into a larger psychometric spread in plots such as Figure 5C. This example was selected to illustrate a region of the MLCRF with shallower slope, indicating relatively lower confidence in the threshold estimate. Conversely, greater certainty leads to narrower transitions. If an application calls for unbiased estimates of psychometric spread, the use of the posterior mean function of a probabilistic classifier observed through a sigmoidal link function has been shown to be effective (Song et al., 2017, 2018). Because of this earlier work and the yet-to-be-established utility of spread for clinical applications of contrast detection, no formal analysis of spread estimation is performed for the current study.

The previous analysis provided proof-of-concept evidence in support of the accuracy and efficiency of the MLCRF estimator and, by extension, the MLCSF estimator. By updating a MLCSF model each time a new data point is collected and comparing to CSF ground truth, the average accuracy of the estimator can be evaluated as a function of the amount of data collected (i.e., the sample count or number of behavioral trials).

In repeat simulations for each canonical phenotype, two independent estimator configurations were evaluated in a 2×2 arrangement: sampling strategy and selection of prior. The two choices for sampling were random, as depicted in Figure 4, and active, as depicted in Figure 5. Random sampling represents a way to quantify the performance of a machine learning estimator independent of its internal optimization algorithm, which can perform poorly for complex functions by failing to converge or becoming trapped at local optima. Assuming the optimizer is functioning well, active sampling is expected to produce systematically better performance than random sampling.

The two choices for prior were uninformative and canonical phenotype mixture. An informative prior incorporates knowledge into the model that originates from beyond the immediately collected data. A prior represents an inductive bias, and a bias in the right direction is expected to improve the efficiency of the estimator, i.e., allowing it to systematically converge to a final estimate faster than with the use of an unbiased, uninformative prior. A poorly chosen bias can, on the other hand, push learning in the wrong direction and decrease efficiency.

The four combinations of these estimator configurations were evaluated for each canonical phenotype 10 times and the resulting average MLCSF accuracy compared against ground truth CSFs plotted as a function of sample count in **Figure 6**. For all canonical phenotypes and estimator configurations, the MLCSF converged toward an accurate representation of ground truth CSF values, as signified by the trend toward low root-mean-square errors. For a visual indication of estimator quality at a given RMSE value, refer to Figures 4 and 5.

**Figure 6:**
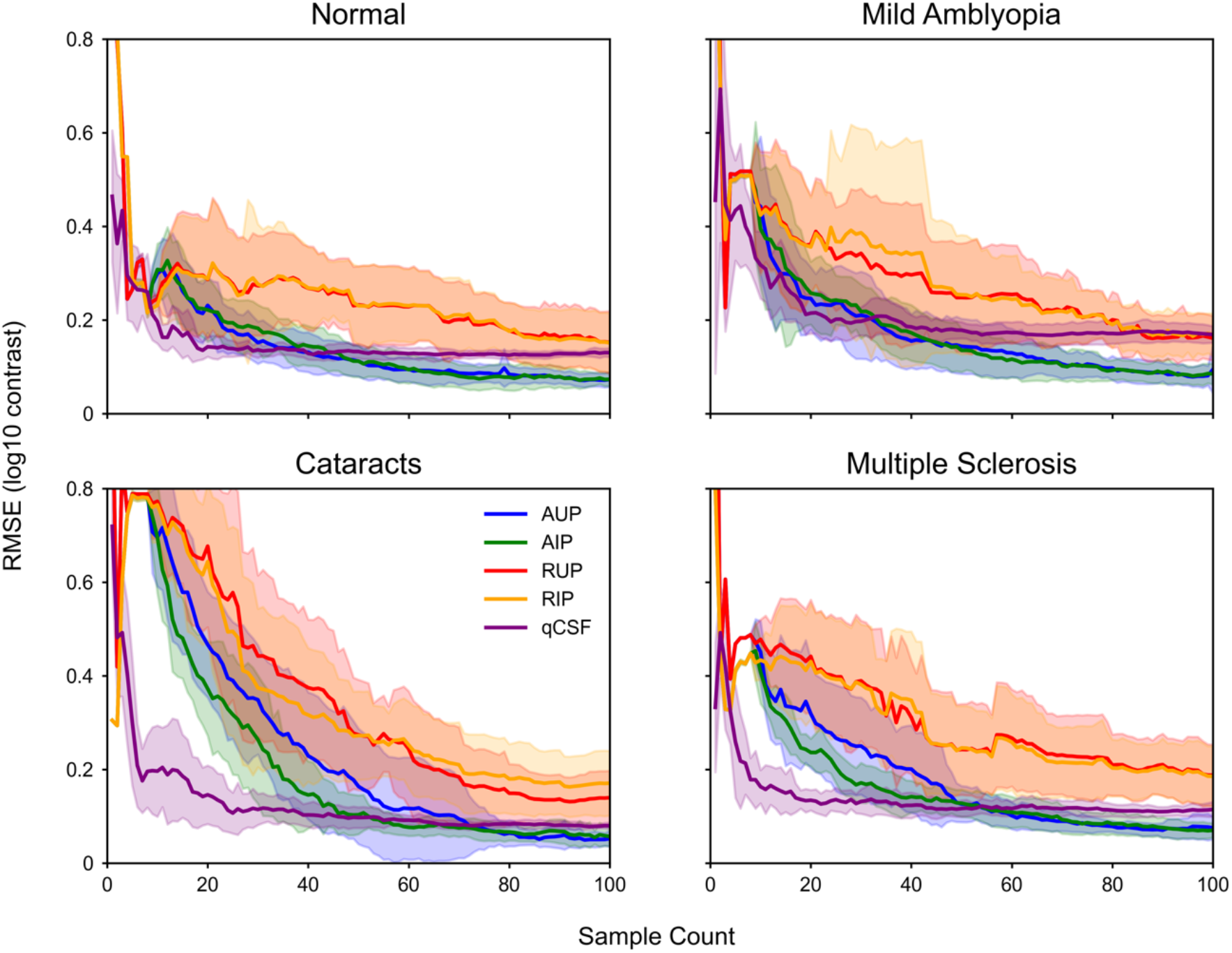
Mean +/– standard deviation RMSE values in log contrast units of all four canonical phenotypes averaged from 10 repeat experiments using up to 100 samples of random with uninformative priors (RUP), random with informative priors (RIP), active with uninformative priors (AUP) and active with informative priors (AIP). Performance of quickCSF on the same estimation tasks is also shown.

As expected, active learning, with sampling intentionally targeting threshold values, accelerated convergence toward accurate CSF representations. Average active sampling performance achieved equivalence to 100-sample average random sampling performance in fewer than 30 samples in all cases. Given that the first 8 samples were deterministic for every run, approximately 20 active samples turned out to be about as effective as approximately 100 random samples. This expected behavior confirms that the optimization component of MLCRF is functioning as designed.

A more complex scenario can be seen, however, when comparing the relative contributions of different priors. Little relative efficiency gain (i.e., more rapid convergence) can be seen with the incorporation of an informative prior, either for random or active sampling. It is possible that any effect of an informative prior at the beginning is muted by the phantom samples and the Halton samples used as a primer sequence. Halton sampling was included based on previous experience to prevent runaway degenerate sampling for the active learning, uninformative prior condition. Perhaps an informative prior can supplant this kind of heuristic, though there is practical value for behavioral testing scenarios in having participants start with a few trials that are very likely to be well above and well below their performance thresholds.

The quickCSF method can be seen to converge to its final estimates even more quickly than active MLCSF, though the final estimates themselves are lower in accuracy. For these canonical phenotypes, quickCSF requires only 20–30 samples to converge. Performance of quickCSF is also consistent across multiple experiments, indicating reliability similar to active MLCSF and considerably higher than random MLCSF.

### Experiment 2

The results of experiment 1 demonstrate that the general form of machine learning psychometric field estimation can successfully be applied to estimating visual contrast response functions for idealized phenotypes. In order to further illustrate the ability of this novel estimator to represent a variety of actual CSFs estimated from individuals in the course of human experiments, the types of manipulations from experiment 1 were replicated for different individuals in a second experiment.

Similarly to experiment 1, each individual CSF depicted in Figure 2 was used to create a generative ground truth model CSF. An example from an individual with schizophrenia plotted according to the conventions of Figure 5 can be seen in **Figure 7**. The flexibility of the machine learning estimator is apparent in this example just as in Figure 5. By happenstance this example includes a lucky guess near (1.5, 0.01), which did not ultimately bias the final threshold estimate, although additional samples in that vicinity were required for the model to confirm that it was an outlier. This detail reveals the robustness of the MLCRF estimator as well as its efficiency.

**Figure 7:**
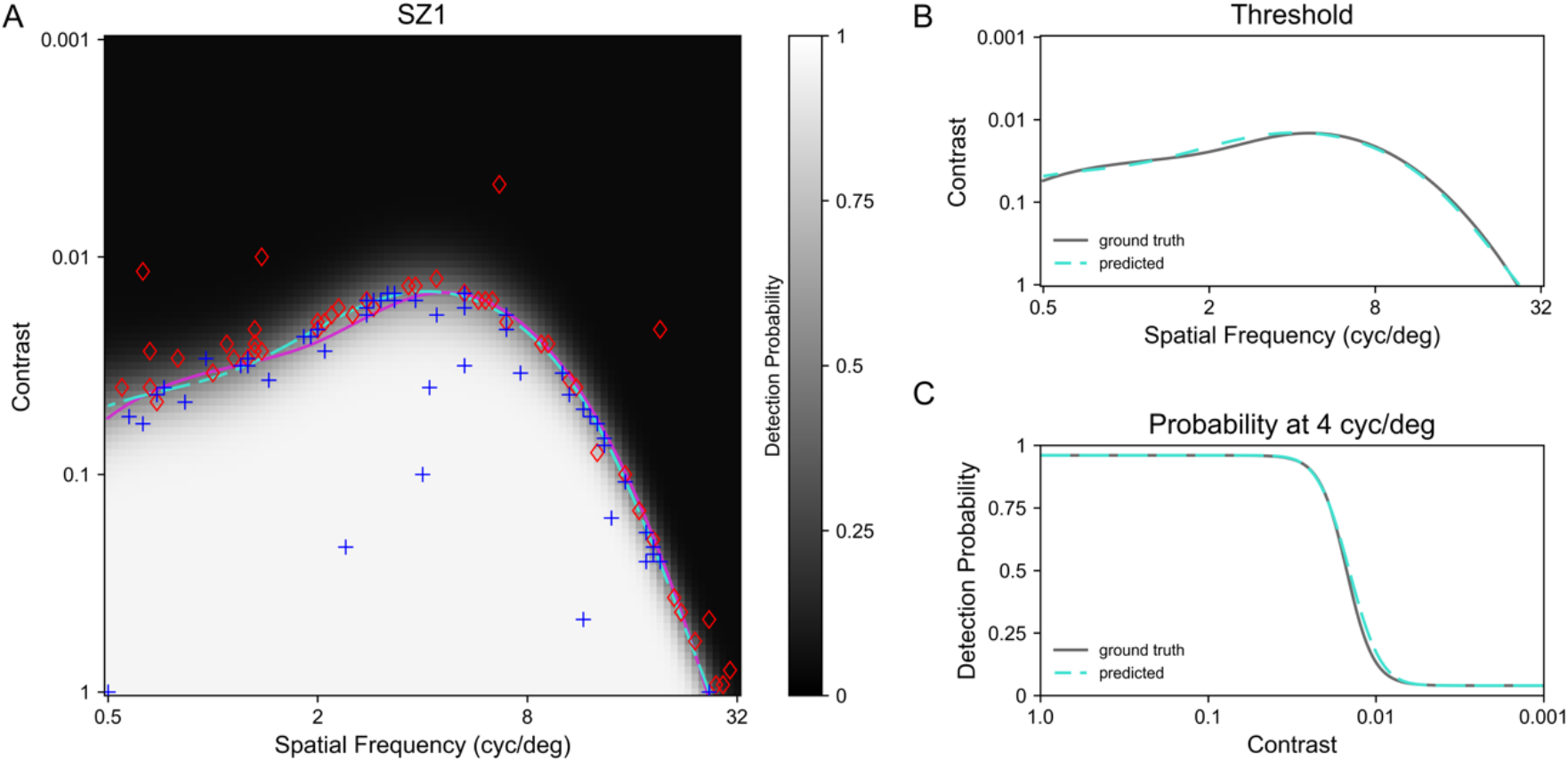
The MLCRF estimated with 100 samples of active learning using plotting conventions of Figure 5. **A**. Simulated behavioral responses, ground truth of a single schizophrenic CSF, and MLCSF estimate superimposed on the predictive posterior MLCRF mean. **B**. MLCSF curve replotted along with the ground truth CSF curve. Ordinate units indicate stimulus contrast. **C**. MLCRF psychometric curve at 4 cycles per degree plotted along with the ground truth psychometric function curve. RMSE value in log contrast units for this MLCSF curve is 0.035.

The individual ground truths compared against MLCSF estimates from 8 Halton + 92 active samples with uninformative priors can be seen in **Figure 8**. Visually, MLCSF estimates show close alignment with the ground truths, also reflected in the low accompanying RMSE values. Notably, several examples deviate substantially from standard parametric forms typically used for CSF estimators, including the truncated parabola. This result further implies that the MLCSF estimator is flexible enough to accurately reflect a wide variety of individual CSF shapes.

**Figure 8:**
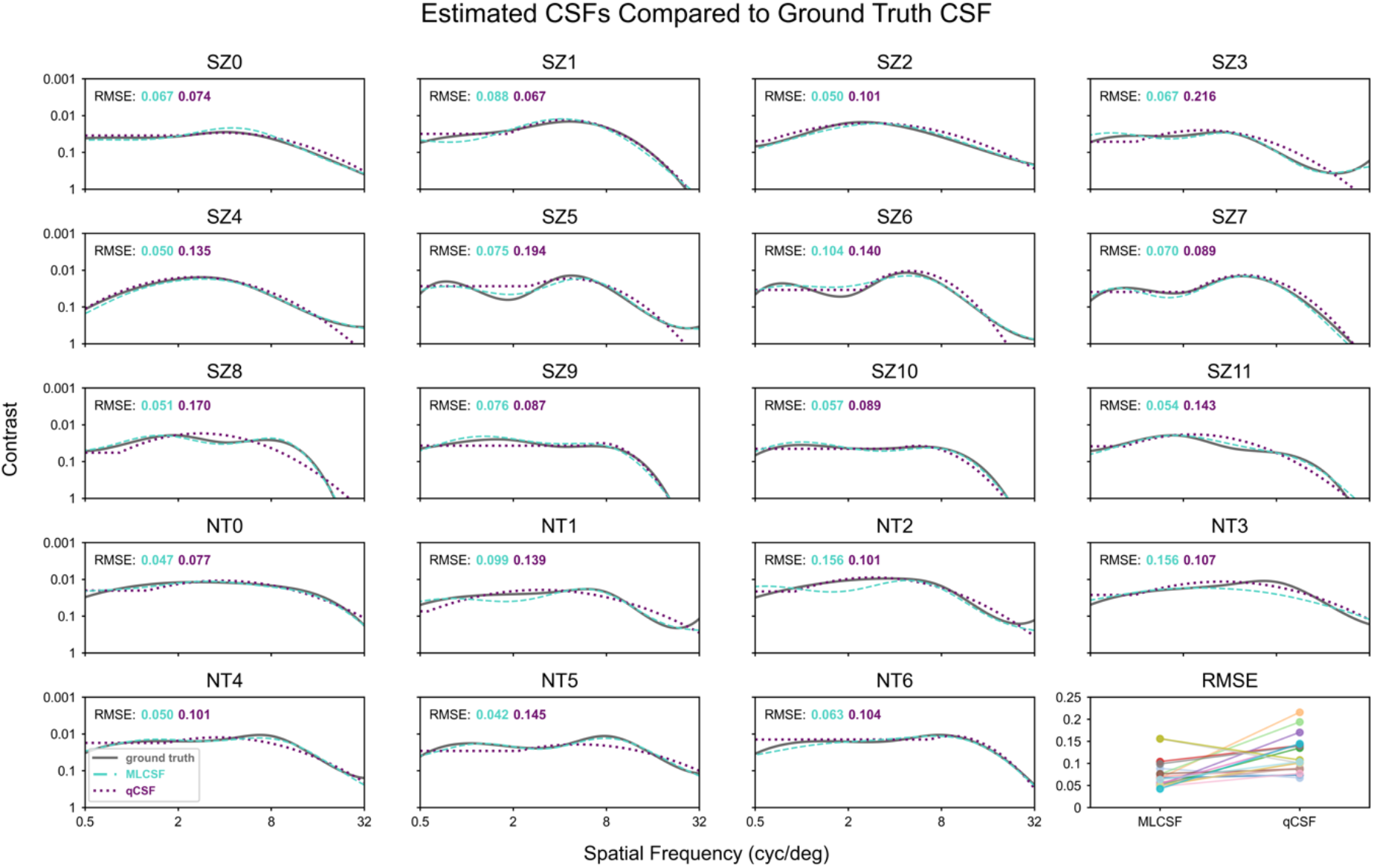
Comparison of ground truth CSF curves with MLCSF and quick CSF curves estimated using 100 active learning samples with uninformative prior. Twelve individuals with schizophrenia and seven neurotypical individuals are represented. Relative accuracy for MLCSF and quickCSF is shown in the lower right panel.

Performance of the quickCSF estimator for this set of ground truths reveals, as expected, that the truncated parabola functional form matches diverse phenotypes with a wide range of goodness-of-fits. The quickCSF RMSE values are almost always larger than the MLCSF values, confirming that the nonparametric estimator is able to fit heterogeneous curves systematically better than a low-order parametric form.

As in experiment 1, the same two independent estimator configurations were evaluated for individuals in a 2×2 arrangement of sampling strategy and prior as a function of sample count and the results shown in **Figure 9**. This time, however, each CSF was modeled only once, and variation in performance was evaluated across the entire cohort. Once again, active learning showed an 80% to 90% efficiency gain in converging to accurate CSF estimates over random sampling. Once again, little systematic effect of the informative prior is observed under these experimental conditions.

**Figure 9:**
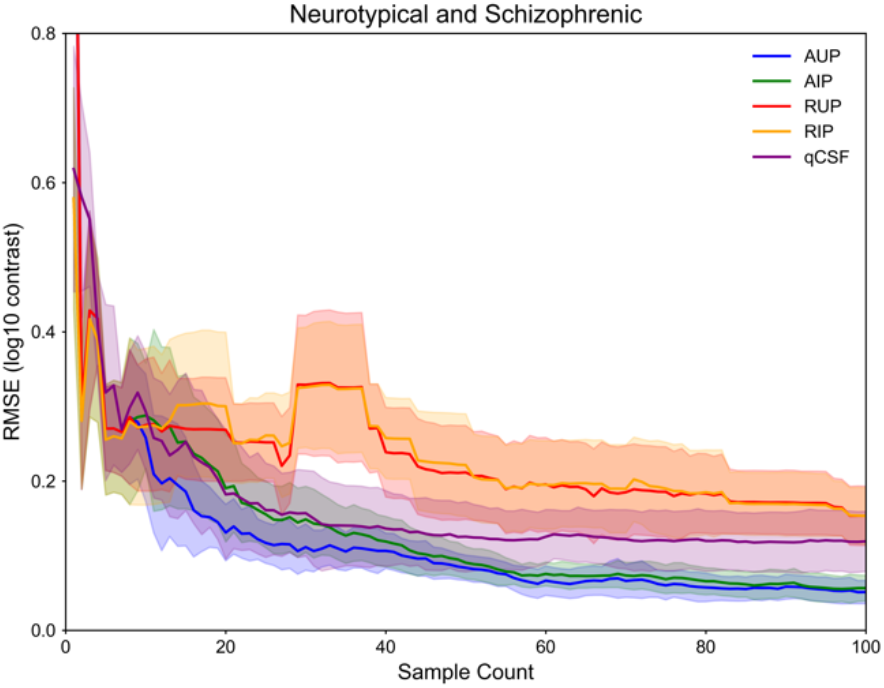
Mean +/– standard deviation RMSE values of all neurotypical and schizophrenic CSFs for the same 4 experimental conditions as in Figure 6, with the same plotting conventions.

Variability in experiment 1 is akin to test-retest reliability of the MLCSF estimator, whereas variability in experiment 2 also includes the variation from different CSF shapes. The latter is therefore likely to be a more appropriate representation of the variation expected in estimator performance for an arbitrary CSF shape. It is also worth observing that variation from active sampling is systematically lower than from random sampling in all conditions, as expected.

Performance of the quickCSF estimator shows some similarity to that observed in Figure 6, with average convergence again completed by around 20–30 samples. Average rate of convergence and average final fit accuracy are lower than active learning MLCSF, and variability is higher. In fact, all quickCSF metrics fare more poorly for experiment 2 than experiment 1. The canonical phenotypes of experiment 1 are smoother than might be typically encountered, while the individual phenotypes recorded from a visual training task could be less smooth than average. While quickCSF shows performance advantages when ground truth phenotypes match its underlying model, the relative lack of flexibility yields more variable performance when phenotypes themselves are more variable.

## Discussion

Psychometric estimator design for research or clinical applications faces common considerations. Nonparametric methods such as adaptive staircases are individually efficient at estimating single thresholds by themselves. When strung together they do produce the most systematically accurate conventional estimators of threshold curves such as the CSF (Watson & Ahumada, 2005), but with diminishing returns for efficiency at high dimensions and resolutions. Figure 2 shows examples of the kinds of curves that result. A major disadvantage of this approach is the need to interpolate threshold values between discretized, pre-selected abscissa values on the curve. For phenotypes with steep dropoffs or other local nuances, this uniform sampling approach decreases estimation accuracy. Furthermore, this discretization limits the ability to exploit additional information to improve estimation efficiency without relying on assumptions about the functional form of the CSF.

Alternatively, parametric methods can incorporate outside information, such as constraints on CSF shape, and provide continuous threshold estimates. CSF estimators designed along these principles can achieve high efficiency at the expense of some accuracy (Hou et al., 2015; Lesmes et al., 2010; Watson & Ahumada, 2005; Zhao et al., 2021). Neither conventional parametric nor nonparametric models of CSF learn item-level prediction, however, because assumptions about the model likelihood are used to enhance efficiency gains.

This paper describes the development of a probabilistic machine learning classifier designed to learn fully predictive models of item-level contrast detection responses from human participants for patterned visual stimuli as a function of spatial frequency and visual contrast. From these machine learning contrast response functions (MLCRFs), machine learning contrast sensitivity functions (MLCSFs) can be estimated as a single performance threshold contour, here taken to be the 0.5 detection probability contour. Similar methods have been successful at estimating detection thresholds for audibility (Cox & de Vries, 2021; Schlittenlacher et al., 2018; Song et al., 2015) and visual fields (Chesley & Barbour, 2020).

As expected for a nonparametric estimator, accuracy for the MLCSF was generally high for a variety of shapes. Efficiency was poor for the tested estimator configuration with random sampling, however, which was not unexpected given previous findings (Song et al., 2018). Active learning led to more informative stimulus delivery, yielding faster convergence to accurate CSF estimates. Including an informative prior did not systematically improve estimation efficiency in these experiments, which was an unexpected result. Perhaps the other elements of the estimator configurations, such as the initial deterministic sampling or the phantom observations, muted the impact of the particular informative prior that was used. A reasonable conclusion of these results is that an informative prior under these conditions provides little performance gain, although it might under other conditions.

Tested on the same ground truth CSFs, the quickCSF estimator lived up to its name and converged rapidly toward its final estimate when ground truth CSFs were stereotypical. In other words, the inductive bias of quickCSF seems to be well matched to canonical phenotypes. QuickCSF performed less well with the phenotypes derived from a perceptual training task, however. In this case the inductive bias of the truncated parabola led to underfits of the resulting phenotypes, leading to an irreversible loss in both accuracy and efficiency. This performance decrement for quickCSF relative to MLCSF occurred even though the former was preferentially advantaged with clean training data, i.e., no losses or guesses to disrupt learning.

The MLCSF inductive bias was not as well matched to canonical phenotypes, leading to slower convergence than quickCSF, but its flexibility allowed it to achieve approximately the same average accuracy on both experiments. Many other options exist to tune the MLCSF inductive bias, including other implementations of CSF priors, mutually conjoint estimation of multiple CSF curves (Barbour et al., 2018; Heisey et al., 2018), multiplexed detection tasks (Gardner, Song, et al., 2015), and perhaps the most efficient of all, Bayesian active model selection from among predefined phenotypes (Gardner, Malkomes, et al., 2015; Larsen et al., 2020, 2021).

In the latter framework, each new data point is used to accumulate evidence for or against discrete hypotheses defined either by phenotypic examples or an individual’s previous test results. This procedure retains the flexibility of nonparametric estimation while requiring quite a small number of samples to reach a confident conclusion. Its potential utility might lie in beginning each eye’s CSF test as a screening procedure to first try to rule out any pathology efficiently. If this cannot be accomplished with high confidence using a small number of samples, the acquisition function could then switch to optimally determining the actual shape of the CSF, as in the current study. At each phase of testing the question of greatest interest is addressed optimally, which leads to considerable time savings for a given sensitivity and specificity. This efficiency advantage is likely to be highest with highly asymmetric pretest probabilities, such as for screening tests in populations with no known visual disorders.

A distinction in clock time also exists between quickCSF and the currently implemented MLCSF. Retraining quickCSF models required a consistent model retraining time of 0.61 ± 0.024s. Retraining MLCSF, however, required steadily increasing model retraining times as data quantity increased (e.g., 1.0 ± 0.019s at 10 data points and 2.3 ± 0.049s at 100 data points). These values allow for real-time model updating compatible with standard pauses between behavioral tests for the typical amounts of data expected for MLCSF. All of these computations were made without GPU acceleration on a Dell Precision 5820 Workstation with Xeon W-2245 8-core 3.90 GHz CPU and 128 GB of RAM.

The MLCRF learns a psychometric spread at every spatial frequency rather than assuming it. Unfortunately, examples of such multidimensional models do not exist to compare against in terms of efficiency. The utility of this capability is unclear at present because spread estimates are not generally employed, but there could be subtle variations in spreads for different phenotypes that might make sensitive testing procedures more likely to resolve certain pathologies (Foley & Legge, 1981). Thus, this fully predictive model could open up avenues in research for the exploration of deeper behavioral response relationships not yet fully considered in psychophysics and vision science.

Finally, while the MLCRF as coded can make use of small or large amounts of data to improve estimation properties, the framework it is based upon could easily accommodate different machine learning algorithms as data available to constrain models increase in number. It is not unreasonable to consider that in the future, once large numbers of individuals have completed MLCRF tests, the mass of data could be used to train even more flexible machine learning models that can exploit many small correlations in widely divergent data streams to accommodate more refined individualized phenotypes. The models themselves could grow in complexity, incorporating more details about visual and brain health than simply a person’s ability to resolve visual images at different spatial frequencies and contrasts.

## Conclusions

A nonparametric Bayesian estimator learning from simulated contrast detection behavioral responses can provide accurate estimates for a variety of contrast sensitivity function shapes in a few dozen samples under ideal conditions. This estimator makes use of recent developments in probabilistic machine learning classification that provide it with potential advantages over classical estimators at the cost of increased model complexity. Notably, it can accommodate a wide set of constraints on functional form; it is a learned predictive model; it is a fully probabilistic model; and it can be extended to incorporate related data from a variety of sources. It is more accurate and, in at least some scenarios, more efficient than classical parametric estimators. It has the potential to become even more efficient by imposing a stronger inductive bias, or more accurate by continuing to learn as more data are acquired. Future work will extend and improve the estimator algorithm in ways described above and evaluate its performance in real-time human data acquisition.

## Data Availability

All data produced in the present study are available at https://osf.io/cpkn5/.

## Acknowledgments

Supported by R21EY033553 and R01EY023582. A patent has been issued covering technology described in this study (Barbour, D. L. et al., 2021).

## References

Barbour, D. L., DiLorenzo, J., Sukesan, K. A., Song, X. D., Chen, J. Y., Degen, E. A., Heisey, K. L., & Garnett, R. (2018). Conjoint psychometric field estimation for bilateral audiometry. Behav Res Meth. 10.3758/s13428-018-1062-3

Barbour, D. L., Song, X., Ledbetter, N., Gardner, J., & Weinberger, K. (2021). Fast, Continuous Psychometric Estimation System Utilizing Machine Learning and Associated Method of Use (United States Patent US11037677B2). https://patents.google.com/patent/US11037677B2

Basha, S. M., & Rajput, D. S. (2019). Survey on evaluating the performance of machine learning algorithms: Past contributions and future roadmap. In Deep Learning and Parallel Computing Environment for Bioengineering Systems (pp. 153–164). Elsevier.

Calderone, D. J., Martinez, A., Zemon, V., Hoptman, M. J., Hu, G., Watkins, J. E., Javitt, D. C., & Butler, P. D. (2013). Comparison of psychophysical, electrophysiological, and fMRI assessment of visual contrast responses in patients with schizophrenia. NeuroImage, 67, 153–162. 10.1016/j.neuroimage.2012.11.019

Canare, D., Ni, R., & Lu, T. (2019). An open-source implementation of the Quick CSF method. Journal of Vision, 19(10), 86b. 10.1167/19.10.86b

Chesley, B., & Barbour, D. L. (2020). Visual field estimation by probabilistic classification. IEEE Journal of Biomedical and Health Informatics, 24(12), 3499–3506. 10.1109/JBHI.2020.2999567

Chung, S. T. L., & Legge, G. E. (2016). Comparing the shape of contrast sensitivity functions for normal and low vision. Investigative Ophthalmology & Visual Science, 57(1), 198–207. 10.1167/iovs.15-18084

Cox, M., & de Vries, B. (2021). Bayesian pure-tone audiometry through active learning under informed priors. Frontiers in Digital Health, 3, 723348. 10.3389/fdgth.2021.723348

Deveau, J., Ozer, D. J., & Seitz, A. R. (2014). Improved vision and on-field performance in baseball through perceptual learning. Current Biology: CB, 24(4), R146–147. 10.1016/j.cub.2014.01.004

DiMattina, C. (2015). Fast adaptive estimation of multidimensional psychometric functions. J Vis, 15(9), 5–5.

Foley, J. M., & Legge, G. E. (1981). Contrast detection and near-threshold discrimination in human vision. Vision Research, 21(7), 1041–1053. 10.1016/0042-6989(81)90009-2

Gardner, J. M., Malkomes, G., Garnett, R., Weinberger, K. Q., Barbour, D., & Cunningham, J. P. (2015). Bayesian active model selection with an application to automated audiometry. Adv Neural Inf Process Syst, 2377–2385.

Gardner, J. M., Pleiss, G., Weinberger, K. Q., Bindel, D., & Wilson, A. G. (2018). Gpytorch: Blackbox matrix-matrix gaussian process inference with gpu acceleration. Advances in Neural Information Processing Systems, 31.

Gardner, J. M., Song, X. D., Cunningham, J. P., Barbour, D. L., & Weinberger, K. Q. (2015). Psychophysical testing with Bayesian active learning. Uncertain Artif Intell, 286–295.

Ginsburg, A. P. (2003). Contrast sensitivity and functional vision. International Ophthalmology Clinics, 43(2), 5–15. 10.1097/00004397-200343020-00004

GPyTorch 1.8.1. (n.d.). Retrieved February 26, 2023, from https://docs.gpytorch.ai/en/stable/

Green, D. M., & Swets, J. A. (1966). Signal Detection Theory and Psychophysics. John Wiley & Sons, Inc.

Gu, H., Kim, W., Hou, F., Lesmes, L. A., Pitt, M. A., Lu, Z.-L., & Myung, J. I. (2016). A hierarchical Bayesian approach to adaptive vision testing: A case study with the contrast sensitivity function. Journal of Vision, 16(6), 15. 10.1167/16.6.15

Halton, J. H. (1964). Algorithm 247: Radical-inverse quasi-random point sequence. Commun ACM, 7(12EDITED), 701–702.

Heisey, K. L., Buchbinder, J. M., & Barbour, D. L. (2018). Concurrent Bilateral Audiometric Inference. Acta Acustica United with Acustica, 104(5), 762–765. 10.3813/AAA.919218

Hensman, J., Matthews, A., & Ghahramani, Z. (2015). Scalable variational Gaussian process classification. Artificial Intelligence and Statistics, 351–360.

Hou, F., Lesmes, L., Bex, P., Dorr, M., & Lu, Z.-L. (2015). Using 10AFC to further improve the efficiency of the quick CSF method. Journal of Vision, 15(9), 2. 10.1167/15.9.2

Houlsby, N., Huszár, F., Ghahramani, Z., & Lengyel, M. (2011). Bayesian active learning for classification and preference learning. ArXiv Preprint ArXiv:1112.5745.

Kalloniatis, M., & Luu, C. (1995). Visual Acuity. In H. Kolb, E. Fernandez, & R. Nelson (Eds.), Webvision: The Organization of the Retina and Visual System. University of Utah Health Sciences Center. http://www.ncbi.nlm.nih.gov/books/NBK11509/

Kingdom, F. A. A., & Prins, N. (2010). Psychophysics: A Practical Introduction. Elsevier.

King-Smith, P. E. (1984). Efficient threshold estimates from yes-no procedures using few (about 10) trials. American Journal of Optometry and Physiological Optics, 61, 119.

King-Smith, P. E., & Rose, D. (1997). Principles of an adaptive method for measuring the slope of the psychometric function. Vision Research, 37(12), 1595–1604. 10.1016/s0042-6989(96)00310-0

Kokol, P., Kokol, M., & Zagoranski, S. (2021). Machine learning on small size samples: A synthetic knowledge synthesis. ArXiv Preprint ArXiv:2103.01002.

Kolb, H., Fernandez, E., & Nelson, R. (Eds.). (1995). Webvision: The Organization of the Retina and Visual System. University of Utah Health Sciences Center. http://www.ncbi.nlm.nih.gov/books/NBK11530/

Kontsevich, L. L., & Tyler, C. W. (1999). Bayesian adaptive estimation of psychometric slope and threshold. Vision Res, 39(16), 2729–2737.

Larsen, T. J., Malkomes, G., & Barbour, D. L. (2020). Accelerating Psychometric Screening Tests With Bayesian Active Differential Selection. ArXiv Preprint ArXiv:2002.01547.

Larsen, T. J., Malkomes, G., & Barbour, D. L. (2021). Accelerating Psychometric Screening Tests with Prior Information. In A. Shaban-Nejad, M. Michalowski, & D. L. Buckeridge (Eds.), Explainable AI in Healthcare and Medicine: Building a Culture of Transparency and Accountability (pp. 305–311). Springer International Publishing. 10.1007/978-3-030-53352-6_29

Leek, M. R. (2001). Adaptive procedures in psychophysical research. Percept Psychophys, 63(8), 1279–1292.

Lesmes, L. A., Jeon, S.-T., Lu, Z.-L., & Dosher, B. A. (2006). Bayesian adaptive estimation of threshold versus contrast external noise functions: The quick TvC method. Vision Research, 46(19), 3160–3176. 10.1016/j.visres.2006.04.022

Lesmes, L. A., Lu, Z.-L., Baek, J., & Albright, T. D. (2010). Bayesian adaptive estimation of the contrast sensitivity function: The quick CSF method. Journal of Vision, 10(3), 17.1-21. 10.1167/10.3.17

Levitt, H. (1971). Transformed up-down methods in psychoacoustics. The Journal of the Acoustical Society of America, 49(2B), 467–477.

PyTorch 1.13. (n.d.). Retrieved February 26, 2023, from https://pytorch.org/docs/stable/index.html

Rasmussen, C. E., & Williams, C. K. I. (2006). Gaussian Processes for Machine Learning. The MIT Press.

Rohaly, A. M., & Owsley, C. (1993). Modeling the contrast-sensitivity functions of older adults. JOSA A, 10(7), 1591–1599. 10.1364/JOSAA.10.001591

Schlittenlacher, J., Turner, R. E., & Moore, B. C. J. (2018). Audiogram estimation using Bayesian active learning. The Journal of the Acoustical Society of America, 144(1), 421. 10.1121/1.5047436

Shams, L., & Seitz, A. R. (2008). Benefits of multisensory learning. Trends in Cognitive Sciences, 12(11), 411–417. 10.1016/j.tics.2008.07.006

Song, X. D., Garnett, R., & Barbour, D. L. (2017). Psychometric function estimation by probabilistic classification. The Journal of the Acoustical Society of America, 141(4), 2513. 10.1121/1.4979594

Song, X. D., Sukesan, K. A., & Barbour, D. L. (2018). Bayesian active probabilistic classification for psychometric field estimation. Attention, Perception & Psychophysics, 80(3), 798–812. 10.3758/s13414-017-1460-0

Song, X. D., Wallace, B. M., Gardner, J. R., Ledbetter, N. M., Weinberger, K. Q., & Barbour, D. L. (2015). Fast, continuous audiogram estimation using machine learning. Ear and Hearing, 36(6), e326–335. 10.1097/AUD.0000000000000186

Tahir, H. J., Parry, N. R. A., Pallikaris, A., & Murray, I. J. (2009). Higher-order aberrations produce orientation-specific notches in the defocused contrast sensitivity function. Journal of Vision, 9(7), 11. 10.1167/9.7.11

Titsias, M. (2009). Variational learning of inducing variables in sparse Gaussian processes. Artificial Intelligence and Statistics, 567–574.

Treutwein, B. (1995). Adaptive psychophysical procedures. Vision Res, 35(17), 2503–2522.

Treutwein, B., & Strasburger, H. (1999). Fitting the psychometric function. Percept Psychophys, 61(1), 87–106.

Wang, X., Wang, H., Huang, J., Zhou, Y., & Tzvetanov, T. (2016). Bayesian Inference of Two-Dimensional Contrast Sensitivity Function from Data Obtained with Classical One-Dimensional Algorithms Is Efficient. Frontiers in Neuroscience, 10, 616. 10.3389/fnins.2016.00616

Watson, A. B., & Ahumada, A. J. (2005). A standard model for foveal detection of spatial contrast. Journal of Vision, 5(9), 717–740. 10.1167/5.9.6

Watson, A. B., & Pelli, D. G. (1983). QUEST: A Bayesian adaptive psychometric method. Perception & Psychophysics, 33(2), 113–120.

Wichmann, F. A., & Hill, N. J. (2001). The psychometric function: I. Fitting, sampling, and goodness of fit. Perception & Psychophysics, 63(8), 1293–1313. 10.3758/BF03194544

Woods, R. L., Bradley, A., & Atchison, D. A. (1996). Consequences of monocular diplopia for the contrast sensitivity function. Vision Research, 36(22), 3587–3596. 10.1016/0042-6989(96)00091-0

Yaghoubi, K. C., Jayakumar, S., Ahmed, A. O., Butler, P. D., Silverstein, S., Thompson, J. L., & Seitz, A. R. (2022). Characterization of training profiles between individuals with schizophrenia and healthy individuals on Contrast Detection and Contour Integration tasks. Journal of Vision, 22(14), 3728–3728.

Yarmohammadi, A., Zangwill, L. M., Diniz-Filho, A., Suh, M. H., Yousefi, S., Saunders, L. J., Belghith, A., Manalastas, P. I., Medeiros, F. A., & Weinreb, R. N. (2016). Relationship between optical coherence tomography angiography vessel density and severity of visual field loss in glaucoma. Ophthalmology, 123(12), 2498–2508.

Zhao, Y., Lesmes, L. A., Hou, F., & Lu, Z.-L. (2021). Hierarchical Bayesian modeling of contrast sensitivity functions in a within-subject design. Journal of Vision, 21(12), 9. 10.1167/jov.21.12.9

Zhou, L., Pan, S., Wang, J., & Vasilakos, A. (2017). Machine Learning on Big Data: Opportunities and Challenges. Neurocomputing, 237, 350–361.

